# Assessment of Safety and Acceptance toward Covid-19 Vaccines: A cross-sectional Study in Egypt

**DOI:** 10.1101/2025.10.28.25338945

**Authors:** Rehab H. Werida, Mohamed T. Amralla, Nader N. Bekhit, Mena Youssef, Gaidaa M. Dogheim

**Author notes:** **Corresponding author: Rehab H. Werida**: Associate Professor and Head of Clinical Pharmacy & Pharmacy Practice Department, Faculty of Pharmacy, Damanhour University, Egypt., **Gaidaa M. Dogheim:** Pharmaceutics Department, Faculty of pharmacy, Alexandria University, Egypt.

## Abstract

**Aim and background:** Coronavirus disease-19 (Covid-19) is a highly spreadable respiratory system disease. Vaccination is a preventive approach to combat the pandemic. The aim of the study was to investigate vaccination pattern in Egypt compared to the world, evaluate the short-term side effects of different vaccines and to assess people’s knowledge, satisfaction, and tolerability of the vaccines.

**Methods:** A cross-sectional, web-based online survey conducted using a Google Form-questionnaire to gather information from the general public regarding their knowledge, satisfaction, and tolerability to COVID-19 vaccine. Demographic data, previous infection with Covid-19, risk factors, type of vaccine received and side effects were collected and statistically analyzed.

**Results:** A total of 508 responses were included in the final analysis. Results showed that most of the participants were females (67.5%), between 20 and 40 years of age (68.3%), and from the medical field (51.1%). The main reported risk factors associated with Covid-19 infection were chronic diseases such as cardiovascular diseases (26.2%), followed by work environment (25%), respiratory conditions (18.2%), and advanced age (16.8%). 57.3% of the respondents received adequate information about the vaccine, 43.3% trusted the information provided while 24.2% required more information. Among all respondents, 78.5% received Covid-19 vaccine were SinoPharm BBIBP was the most administered vaccine (30.4%). Reported adverse effects were injection site pain (18.3%), headache (17.9%), fever (16.8%), fatigue (20.9%), muscle aches (15.1%), nausea (10.8%) and menstrual irregularities (0.2%). Seriousness of Covid-19 infection was the main reason for accepting Covid-19 vaccination (69.7%), followed by believing in the safety and efficacy of the vaccine (36%), while travel was reported with a percentage of 22.2%.

**Conclusion:** The study showed that the participants generally had a good perception and acceptance of Covid-19 vaccination. The main adverse events reported were fever, fatigue and headache. Age is an independent profound variable influencing tolerability of Covid-19 vaccine.

**Impact of findings on practice statements:**

- Good perception and acceptance of Covid-19 vaccination, depend on the factual information from trusted sources, mainly the ministry of health.
- Age significantly influenced tolerability of Covid-19 vaccine among participants.
- Demographic factors impact how people view and perceive Covid-19 vaccinations.

## Introduction

Coronavirus disease 2019 (COVID-19) has spread over the world, since its discovery in Wuhan, China in December 2019, and the World Health Organization (WHO) proclaimed it a pandemic on March 11, 2020 [1]. By January 2023, Covid-19 was the pandemic of 2021-2022 and had resulted in more than 664 million confirmed cases and more than 6 million fatalities worldwide [2]. Covid-19 symptoms, according to recent research, can vary from moderate respiratory disease producing fever, dry cough, dyspnea, myalgia, and tiredness to more severe manifestations of pneumonia, cardiac problems necessitating intensive care unit (ICU) hospitalization, and mechanical ventilation [3]. Since the outbreak, medical efforts were channeled in order to manage and treat Covid-19. Different protocols have been implemented in order to contain the pandemic including several therapeutic modalities. These includes antiviral drugs such as favipravir, remdesivir, corticosteroids, immuno-modulatory agents, and monoclonal antibodies such as casirivimab/imdevimab and bamlanivimab [4]. Unfortunately, several variants have developed over the past 2 years, each possess different spectrum of signs and symptoms. Among these variants are beta, gamma, delta, epsilon, and omicron [5].

The delta variant was classified as a variant of concern (VOC) by the WHO on June 15, 2021 then classified as variants being monitored (VBM) on April 14, 2022 [5]. While omicron variant was classified as VOC on November 26^th^ 20221 [5, 6]. As a result, controlling the spread of the virus through the development of vaccines has been undertaken. Several vaccines have been developed, four of which were FDA approved for emergency use which are the Pfizer-BioNTech, Moderna, Johnson & Johnson, and Novavax vaccines [7]. The Pfizer-BioNTech and Moderna vaccines are mRNA vaccines with efficacy of 95% and 94.1%, respectively [8, 9]. The johnson & johnson vaccine is viral vector vaccine with an efficacy of 66.9% at 14 days and 66.1 at 28 days in moderate to severe infections while in severe infections, the efficacy was 76.7% and 85.4% at day 14 and 28 days, respectively [10]. The Novavax vaccine is an adjuvanted vaccine containing SARS-CoV-2 spike protein and Matrix-M adjuvant with an efficacy of 89.7% [11]. Other vaccines developed include AstraZeneca, Sinopharm, Sputnik, and Sinovac vaccines. Reported adverse effects due to the vaccines range from mild to moderate such as fever, chills, headache, fatigue, vomiting, diarrhea muscle aches and pain at the injection site [12]. The frequency of serious adverse events is typically low and include anaphylaxis, myocarditis and pericarditis, acute myocardial infarction and thrombosis [13]. The incidence of adverse effects is significantly lower among inactivated vaccines, protein subunit vaccines, and DNA vaccines as compared to RNA vaccines, non-replicating vector vaccines, and virus-like particle vaccines [14]. The “Health Belief Model” is a theoretical model which can understand and predict the people’s acceptance of the vaccines which can be affected not only by perceived benefits such as effectiveness but also barriers such as safety [15]. In general, the acceptance of the vaccines depends on several parameters such as availability, convenience of access and people’s confidence in vaccination in addition to socioeconomic factors like age, gender, and income [16, 17]. There are worldwide variations in the acceptance of Covid-19 vaccines especially as the vaccines distribution is uneven around the world [18]. Ecuador, Malaysia, Indonesia, and China had the greatest acceptance rates for the Covid-19 vaccination (97.0%, 94.3%, 93.3%, and 91.3% respectively) [19]. On the other hand, Kuwait, Jordan, Italy, Russia, Poland, the United States, and France had the lowest rates of acceptance (23.6%, 28.4%, 53.7%, 54.9%, 56.3%, 56.9%, and 58.9%, respectively) [19]. In China, vaccine acceptance rate was high especially among healthcare workers [20, 21]. While in the USA, high acceptance rates were observed among the elderly and those with high education and income [22]. According to recent study by *Shawki et al.*, [23] vaccine acceptance rate in Egypt is low where only 14% received or registered to be vaccinated. Furthermore, a number of studies showed that males had higher vaccination acceptance than females [19]. In addition, social media had a great impact on influencing the acceptance of Covid-19 vaccines. In a cross-sectional study, results showed that social media increased the awareness of the vaccines and led to increased acceptance of the vaccines [24]. However, some information about the vaccines were misleading and had a negative effect on people’s acceptance [24]. Thus, social media should share up-to date scientific information about the vaccines to increase the awareness of people about the vaccines [24]. Yet, consulting healthcare professional for any inquiries remains the best approach to obtain evidence-based information [24]. A common cause of vaccine hesitancy is the fear of side effects in addition to availability and doubts in efficacy. Thus, determining the motives behind the vaccine hesitancy across different socioeconomic groups can help us develop policies to reach a population coverage high enough to develop herd immunity [25]. The primary aim of this study was to investigate Covid-19 vaccination patterns worldwide as compared to Egypt. Secondary aim was to determine the short-term side effects of different vaccines available in Egypt and understand people’s perception and acceptance of the vaccines and determine factors which influenced their decision.

## Methods

### Analysis of Vaccination Patterns

To understand the global vaccination pattern for Covid-19 and determine Egypt’s situation, datasets from “COVID-19 Data Explorer” (*Edouard Mathieu, Hannah Ritchie, Lucas Rodés-Guirao, Cameron Appel, Daniel Gavrilov, Charlie Giattino, Joe Hasell, Bobbie Macdonald, Saloni Dattani, Diana Beltekian, Esteban Ortiz-Ospina, and Max Roser (2020) - “Coronavirus (COVID-19) Vaccinations” Published online at* OurWorldinData.org*. Retrieved from: ’*https://ourworldindata.org/covid-vaccinations*’ [Online Resource]*) were retrieved and analyzed using R (version 4.4.3) and R studio software (version 2024.12.1), mainly ‘ggplot2’ and ‘dplyr’ packages. The datasets analyzed were 1) Percentage of people receiving at least one dose of the vaccine, 2) Percentage of people receiving full vaccine protocol, 3) Covid-19 vaccine doses administered per 100 people, by income group, and 4) cumulative Covid-19 doses administered per 100 people (World vs. Egypt).

### Study Design

A cross-sectional study was conducted over the period from December 2021 to February 2022. The subjects participating in the study were residents in Egypt. An online questionnaire, designed on Google Forms, was written in Arabic language and delivered to participants via social media (primarily Facebook, WhatsApp and Linkedin). The participants were not given any incentives to participate. Yet, the importance to fill the forms to conduct this research was clarified. All participants informed at beginning of the questionnaire that information provided about the study have been read and understood, and that filling the questionnaire, indicates participation in this study voluntary. This study was conducted in accordance with the declaration of Helsinki. The settings were adjusted to that each participant sent only one response. Responses were carefully reviewed to ensure that they were not repeated.

The questionnaire comprised two categories of inquiries (**Supplementary Table 1**). The first category covered demographic data of the subject, such as age, gender, educational level, residency, occupation and previous infection with SARS-CoV-2. The second category covered questions about Covid-19 vaccine-related data. These included whether they had taken the vaccine or not, the type of vaccine administered, side effects encountered from the vaccine (headache, fever, fatigue, pain at site of injection and menstrual irregularities), if they got enough information regarding the vaccine and where did they obtain such information from and if they were reluctant to taking the vaccine and the reasons for their hesitancy. All participants were permitted to terminate the survey at any time of the study. All precautionary actions were applied to preserve data confidentiality. The questionnaire was pre-tested for validity. First two experts assessed all the questions individually and minor modifications were made based on their feedback.

### Study collected data

Age, gender, occupation, and academic degree, previous infection with Covid-19, risk factors for Covid-19 infection, type of vaccine received and side effects encountered.

### Sample Size

The minimum sample size for conducting this survey was (500) with a 5% margin of error and a 95% confidence level. In this study, 1,635 subjects participated in the Covid-19 vaccine questionnaire. The number of participants who correctly filled and completed the questionnaire was 508.

### Ethical Consideration

The survey was voluntary and anonymous, and respondents were advised that they might withdraw at any moment. Prior to commencing the questionnaire, informed consent was sought electronically using the form (participants informed about the study at the beginning of the questionnaire and that all information provided will be kept confidential and will be used only for research purposes and participation in this research is entirely voluntary). This study was registered on ClinicalTrial.gov with code no.: NCT05857176. Institutional review board (IRB) approval is deemed unnecessary for online survey according to national regulations.

### Statistical Analysis

Statistical analysis was performed using SPSS version 26.0 (SPSS Inc, Chicago, IL, USA) and R version 4.4.3. Categorical variables were represented as counts and percentages. Chi square test of independence was used to assess bivariate association between categorical variables. Multi-nominal logistic analysis was performed to measure the probability of occurrence of adverse events according to different vaccines. Significance level was set at P < 0.05.

## Results

### Vaccination Patterns Worldwide versus Egypt

Covid-19 vaccination pattern were investigated to understand the global perception and acceptance of the vaccine. The top 50 countries with the highest percentage of people receiving at least one dose (**Figure 1B**) and full vaccination protocol **(Figure 1C**) show a high percentage > ∼70% reflecting acceptance of the vaccine. To understand the influence of income status on vaccine acceptance, as shown in **Figure 1D**, high- and upper-middle income countries have higher percentage of vaccinated people as compared to lower-middle and low income countries. As compared to the world, Egypt has significantly lower percentage of people vaccinated (P < 0.0001) **(Figure 1E)** which reflect lower acceptance and perception of the vaccine.

**Figure 1:**
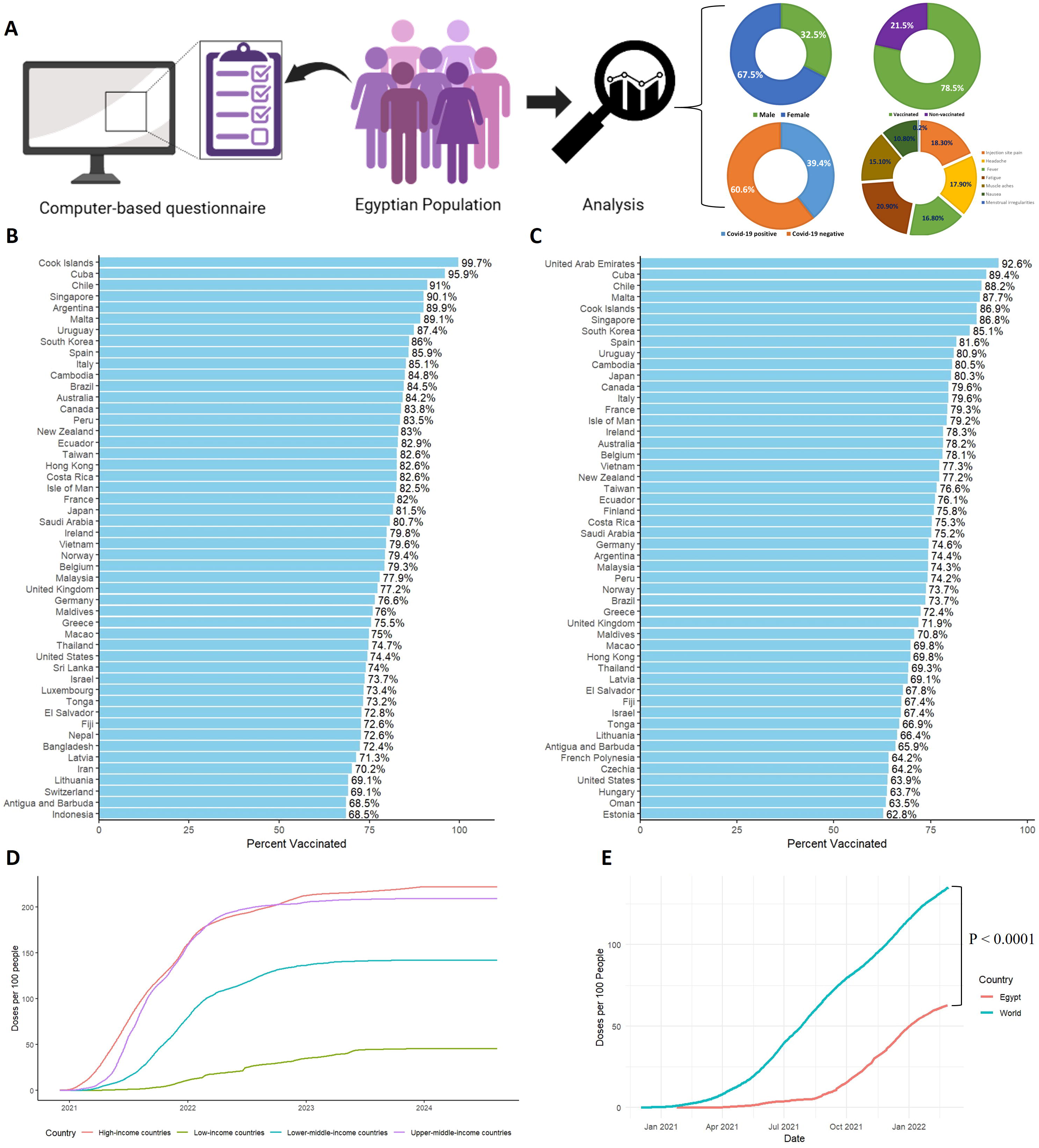
**A**: Schematic presentation of the cross-sectional observational study conducted on Egyptian population. **B**: Percentage of people vaccinated at least one dose of Covid-19 vaccine showing the top 50 countries. **C**: Percentage of people vaccinated the full protocol of Covid-19 vaccine showing the top 50 countries. **D**: Percentage of total people vaccinated grouped by “Income status”. **E**: Percentage of people vaccinated worldwide versus Egypt (till 28-02-2022) showing significant difference at P < 0.0001.

### The Study Participants’ Demographic Characteristics

**Figure 2A** demonstrates the flow diagram of included participants in the online questionnaire. A total of 1,635 responses were submitted, out of which, 472 records didn’t meet eligibility criteria. Then, 1,481 records were screened for data collection where 850 records were excluded due to missing data. Initially 631 were included in the analysis among which 123 records were considered biased and a final of 508 records were included in the final analysis. **Table 1** illustrates the demographic data of participants. The participants were of different age groups as follow: less than 20 years (9.3%), 20 to 30 years (36%), 31 to 40 years (32.3%), 41 to 50 years (13%), and more than 50 years (9.4%). Percentage of female respondents was higher than males (67.5% versus 32.5%). Participants were from Delta region, Egypt. Most respondents were from the healthcare setting (nurses, pharmacists and physician) with a total percentage of 51% and dentists (8.5%), while non-healthcare respondents included accountants (6.7%), bankers (4.9%), engineers (6.7%), housewives (4.5%), marketing and sales (2.4%), teachers (3.5%), and students (11.6%). Contact with Covid-19 patients was investigated, 56.5% of respondents had direct contact with Covid-19 patients. Fifty percent had contact with patients in isolation hospitals, 26% in pharmacies and 24% in clinics.

**Figure 2:**
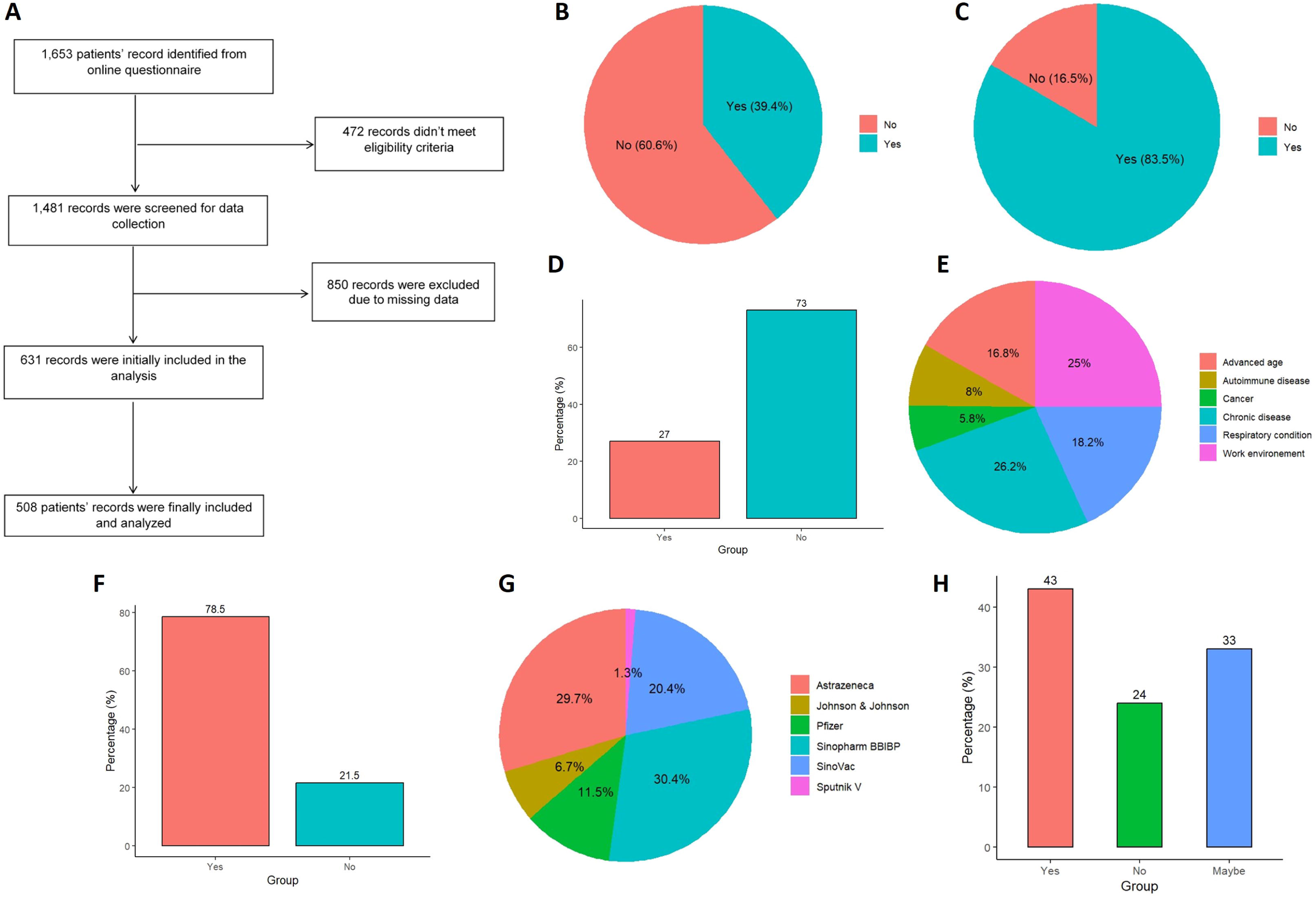
Results of the analyzed data from questionnaire responses. **A**: STROBE Flow Diagram for included Participants. **B**: Percentage of respondents diagnosed with Covid-19. **C**: Percentage of respondents that believe in the seriousness of Covid-19 infection. **D**: Percentage of respondents having risk factors during Covid-19 pandemic. **E**: Risk factors reported by respondents which might increase likelihood of infection. **F**: Percentage of respondents receiving Covid-19 vaccine. **G:** Percentage of different vaccines grouped by manufacturer available in Egypt. **H**: Percentage of respondents who will didn’t receive the vaccine yet but agree to take it.

**Table 1:**
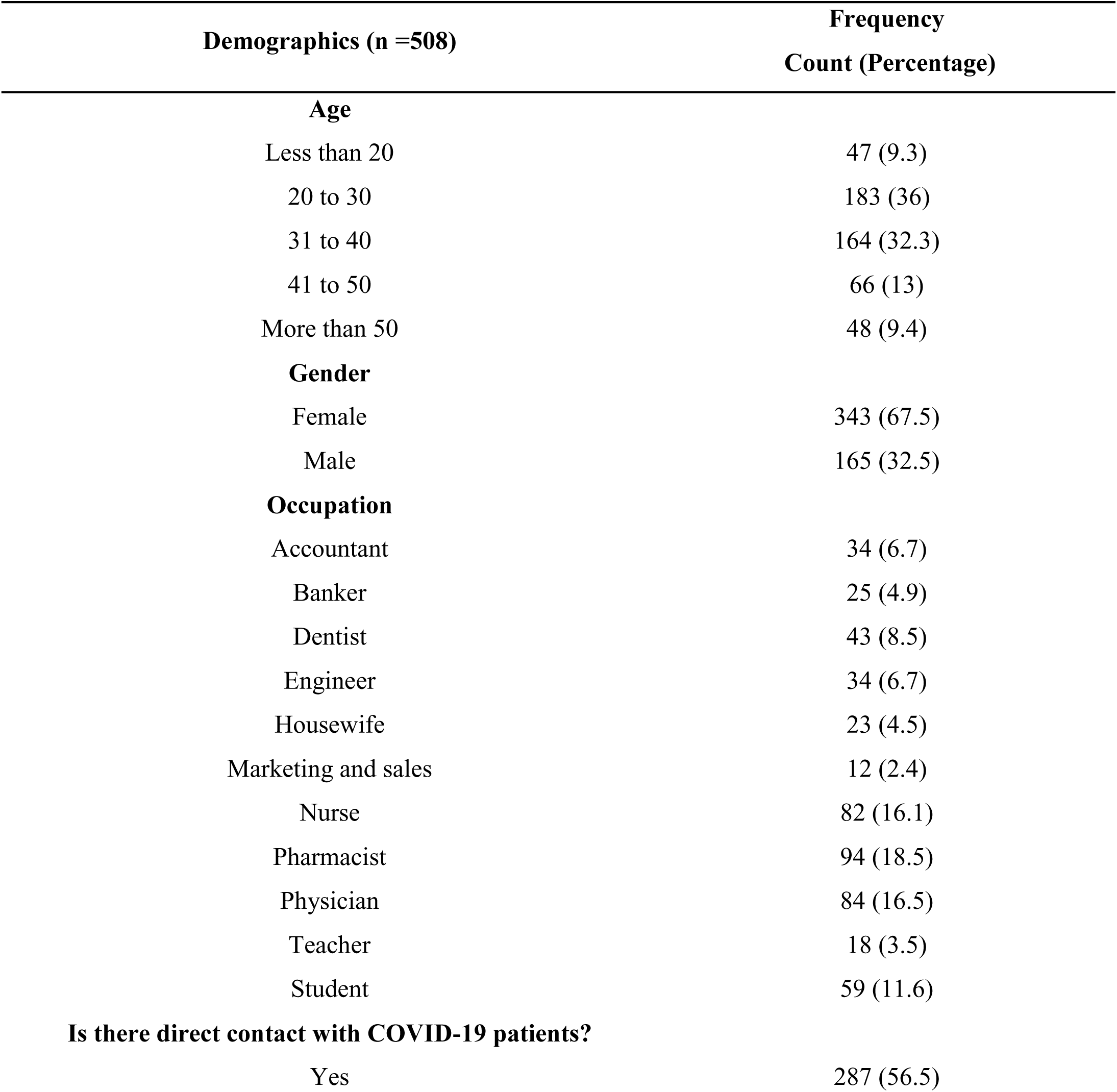
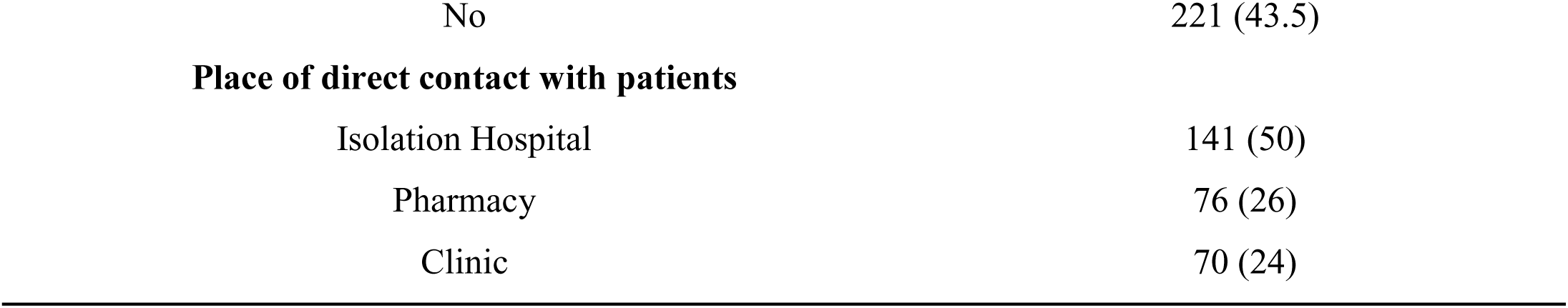
Demographic Data of the Study Participants:

### Covid-19 infection, Risk factors and People’s Perceptions

Percentage of people diagnosed with Covid-19 infection is illustrated in Figure 2B. The majority of respondents reported that they haven’t been diagnosed with Covid-19 infection with a percentage of 60.6%. Perception of participants of the seriousness of Covid-19 infection was investigated. It was found that 83.5% of respondents believe that Covid-19 is a serious infection (Figure 2C). Risk factors implication with the increased risk of Covid-19 infection was also assessed. Twenty seven percent of respondents had risk factors which increase the risk of Covid-19 infection (Figure 2D). Then, the percentage of different risk factors in this group was determined. The majority reported chronic diseases such as cardiovascular diseases as their risk factor (26.2%), followed by work environment (25%), respiratory conditions (18.2%), advanced age (16.8%), autoimmune diseases (8%) and cancer (5.8%) (Figure 2E).

### Covid-19 Vaccination and Types of Vaccines Available in Egypt

The percentage of people who received the vaccine is illustrated in Figure 2F. Among the 508 respondents, 399 people (78.5%) received the vaccine. The different types of vaccines are shown in Figure 2G. SinoPharm BBIBP was the most administered vaccine with a percentage of 30.4%, followed by AstraZeneca (29.7%), SinoVac (20.4%), Pfizer (11.5%), Johnson and Johnson (6.7%), and Sputnik (1.3%). Respondents who didn’t receive the vaccine were further asked about their willingness to receive the vaccine later (Figure 2H). Forty three percent reported their agreement to receive the vaccine, and 33% were still hesitant about the vaccine.

### Participants’ Perception of Covid-19 Vaccines

**Figure 3A** illustrates the questions asked to understand people’s perception of Covid-19 vaccines. First, we investigated the percent of participants believing in the safety and effectiveness of the vaccines, 45.7% of respondents responded with “yes”. Then, we assessed how much information about the vaccines was received and to what extent did they trust this information. The majority received adequate information about the vaccine (57.3%), 43.3% trusted the information provided while 24.2% required more information. Workplace advice which might have influenced people’s perception have been studies as well. The majority of people indeed received advice from their workplace with a percentage of 73.8%. Lastly, we asked the participants if they will advise others to receive the vaccine. A percentage of 79.9 reported that they will advise people to receive the vaccine.

**Figure 3:**
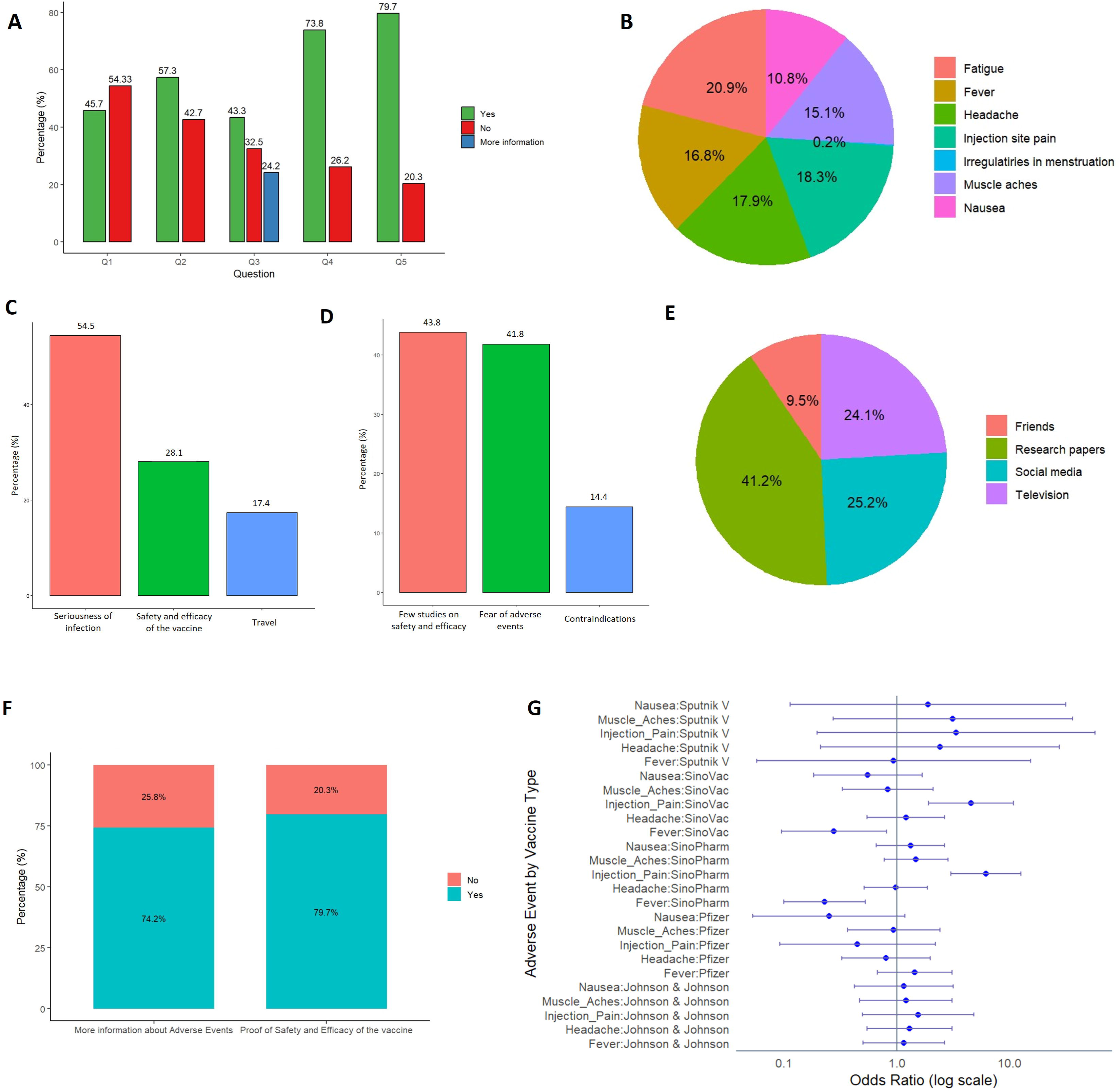
Results of the analyzed data from questionnaire responses. **A:** Different questions that aim at understanding respondents’ perception of Covid-19 vaccine, (Q1: Do you believe in the safety and effectiveness of the vaccines?, Q2: Did you receive adequate information about the vaccine?, Q3: Do you trust the information provided about the vaccine?, Q4: Was their influence from your workplace to receive the vaccine?, Q5: Will you advice other people to receive the vaccine?). **B**: Percentage of adverse events reported by the respondents who received covid-19 vaccine. **C**: Reasons behind the acceptance of covid-19 vaccine include believing in the seriousness of Covid-19 infection, believing in the safety and efficacy of the vaccine and travel purposes. **D:** Reasons behind hesitation or refusal to receive the vaccine which include lack of data on safety and efficacy, fear of adverse events or contraindications to the vaccine. **E:** The impact of social media on the acceptance of the vaccine as compared to other influence factors. **F:** Opinion of respondents on how to increase their acceptance to the vaccine. **G**: Forest plot showing the probability of adverse events occurrence by vaccine type.

### Safety and Reported Adverse Effects due to Covid-19 Vaccines

**Figure 3B** illustrates different adverse effects encountered after the administration of Covid-19 vaccines. The reported adverse effects were injection site pain (18.3%), headache (17.9%), fever (16.8%), fatigue (20.9%), muscle aches (15.1%), nausea (10.8%) and menstrual irregularities (0.2%). We further investigated the percentage of the reported adverse events from each vaccine as shown in **Table 2**. The two most common adverse effects with AstraZeneca vaccine were fever (42.01%) and fatigue (39.5%). Similarly, adverse effects due to Johnson & Johnson vaccine were fever (59.26%), headache (51.85%) and fatigue (48.15%). Participants who received Pfizer vaccines reported fever to be the most common side effect (50%), while injection site pain and nausea were the least reported adverse effects with a percentage of 4.35 each. Sinopharm and Sinovac vaccines had similar adverse effects profile with injection site pain being the most common side effect (55.24% and 28.5%, respectively). The most common adverse effects associated with Sputnik V vaccine were headaches (40%) and muscle aches (40%). Nausea was reported as an adverse effect with all vaccines with the following percentages: AstraZeneca (21.55%), Johnson & Johnson (29.63%), Pfizer (4.35), Sinopharm (21.31), Sinovac (6.1%) and Sputnik V (20%). Incidence of menstrual irregularities was rare, reported only with AstraZeneca with a percentage of 0.84.

**Table 2:** Adverse Effects Reported by Participants after Receiving Different Types of Vaccines.

| Vaccine | Reported Adverse Effects (n = 399) |  |  |  |  |  |  |
| --- | --- | --- | --- | --- | --- | --- | --- |
|  | Injection Site Pain | Headache | Fever | Fatigue | Muscle aches | Nausea | Irregularities in menstruation |
| **AstraZeneca (n = 119)** | 14 (11.76) | 39 (32.77) | 50 (42.01) | 47 (39.50) | 30 (25.2) | 25 (21.55) | 1 (0.84) |
| **Johnson & Johnson (n = 27)** | 6 (22.22) | 14 (51.85) | 16 (59.26) | 13 (48.15) | 10 (37.03) | 8 (29.63) | 0 |
| **Pfizer (n = 46)** | 2 (4.35) | 10 (21.74) | 23 (50.0) | 15 (32.61) | 9 (19.66) | 2 (4.35) | 0 |
| **SinoPharm (n = 122)** | 68 (55.24) | 30 (24.59) | 9 (7.38) | 37 (30.32) | 35 (26.69) | 26 (21.31) | 0 |
| **SinoVac (n = 82)** | 23 (28.05) | 17 (20.73) | 5 (6.10) | 17 (20.73) | 9 (23.17) | 5 (6.10) | 0 |
| **Sputnik V (n = 5)** | 1 (20) | 2 (40) | 1 (20) | 1 (20) | 2 (40) | 1 (20) | 0 |

### Vaccine Hesitancy and Factors Affecting Acceptance of Vaccines

Reasons behind the acceptance of Covid-19 vaccine were investigated (Figure 3C). Seriousness of Covid-19 infection was the main reason with a percentage of 69.7%, followed by believing in the safety and efficacy of the vaccine with a percentage of 36%, while travel was reported with a percentage of 22.2%. Moreover, reasons behind their hesitation or refusal to receive the vaccine were assessed (Figure 3D). The main reasons were the lack of sufficient studies that provide evidence of safety and effectiveness (64%) and fear of adverse events (61%), while those having contraindication to the vaccines were only 21% of all respondents. The impact of social media on the acceptance of the vaccine was studied by identifying the resources used by participants to obtain information about the vaccines (Figure 3E). Research papers were the main source of Covid-19 information resource with a percentage of 36.8%. Television and social media were also identified as sources of information with a percentage of 21.5 and 22.5, respectively while friends constituted only 8.5%. To overcome the vaccine hesitancy, we asked the participants whether providing more information about the adverse effects and the proof of vaccine safety and efficacy might increase their acceptance of the vaccine (Figure 3F). The majority of participants agreed that more information about adverse events would indeed increase vaccine acceptability with a percentage of 74.2%. In addition, 79.7% of the respondents reported that a proof of safety and efficacy of the vaccine might improve its acceptance.

### Probability of Adverse event occurrence according to vaccine type

**Figure 3G** shows the probability of the occurrence of the reported adverse events according to the vaccine type. Fever occurred at a significantly higher probability in participants who were administered SinoPharm and SinoVac vaccine (OR= 0.23, CI (0.1–0.52), P = 0.000, OR= 0.28, CI (0.09–0.81), P = 0.019, respectively) **(Table 3)**. Similarly, injection-site pain was mostly reported after both SinoPharm and SinoVac vaccine administration (OR= 6.71, CI (3.01–12.66), P = 0.000, OR= 4.45, CI (1.91–10.79), P = 0.001, respectively) **(Table 3)**.

**Table 3:** Probability of Adverse Events occurrence according to Vaccine Type.

| Adverse Event | Vaccine | Odds Ratio | 95% CI | p-value |
| --- | --- | --- | --- | --- |
| Fever | Johnson & Johnson | 1.16 | (0.5–2.66) | 0.732 |
|  | Pfizer | 1.44 | (0.67–3.09) | 0.348 |
|  | SinoPharm | 0.23 | (0.1–0.52) | <0.0001**** |
|  | SinoVac | 0.28 | (0.09–0.81) | 0.019* |
|  | Sputnik V | 0.94 | (0.06–15.46) | 0.965 |
| Headache | Johnson & Johnson | 1.30 | (0.55–3.09) | 0.555 |
|  | Pfizer | 0.80 | (0.32–1.99) | 0.636 |
|  | SinoPharm | 0.98 | (0.51–1.86) | 0.944 |
|  | SinoVac | 1.21 | (0.54–2.67) | 0.646 |
|  | Sputnik V | 2.41 | (0.21–27.59) | 0.479 |
| Injection-site pain | Johnson & Johnson | 1.55 | (0.5–4.83) | 0.450 |
|  | Pfizer | 0.45 | (0.09–2.2) | 0.322 |
|  | SinoPharm | 6.17 | (3.01–12.66) | <0.0001**** |
|  | SinoVac | 4.54 | (1.91–10.79) | 0.001*** |
|  | Sputnik V | 3.36 | (0.2–57.18) | 0.403 |
| Muscle aches | Johnson & Johnson | 1.21 | (0.47–3.09) | 0.698 |
|  | Pfizer | 0.94 | (0.37–2.42) | 0.898 |
|  | SinoPharm | 1.48 | (0.77–2.84) | 0.236 |
|  | SinoVac | 0.83 | (0.33–2.1) | 0.693 |
|  | Sputnik V | 3.13 | (0.27–36.08) | 0.360 |
| Nausea | Johnson & Johnson | 1.16 | (0.42–3.16) | 0.776 |
|  | Pfizer | 0.25 | (0.05–1.18) | 0.081 |
|  | SinoPharm | 1.32 | (0.66–2.65) | 0.434 |
|  | SinoVac | 0.55 | (0.18–1.68) | 0.295 |
|  | Sputnik V | 1.88 | (0.11–31.35) | 0.660 |

Multinomial Logistic Regression at significance level of \*P < 0.05, \*\* P < 0.01, \*\*\* P < 0.001, \*\*\*\* P < 0.0001.

### Correlation Studies

The study further investigated whether age or gender directly correlate with Covid-19 infection. As shown in **Table 4a**, there was no significant correlation between either age group or gender with Covid-19 infection. **Table 4b** demonstrates the lack of association between previous Covid-19 infection and the increased incidence of adverse events after receiving the vaccine. The age group between 20 to 30 years had a significantly higher probability to suffer from injection-site pain (P = 0.000), headache (P = 0.000), fever (P = 0.000), fatigue (P = 0.001) and muscle aches (P = 0.002) as compared to other age groups. On the other hand, there was no significant difference between males and females in the incidence of adverse events **Table 4c**.

**Table 4a:** Correlation between Age/Gender and Infection with COVID-19.

| Parameter | Previous infection with Covid-19 |  |  | P-value |
| --- | --- | --- | --- | --- |
|  | Yes<br>(count/percentage) | No<br>(count/percentage) | Total<br>(count/percentage) |  |
| Injection-site pain | 50/25 | 150/75 | 200/100 | 0.265 |
| Headache | 51/25.5 | 149/74.5 |  | 0.130 |
| Fever | 44/22 | 156/78 |  | 0.492 |
| Fatigue | 52/26 | 148/74 |  | 0.865 |
| Muscle aches | 42/21 | 158/79 |  | 0.243 |
| Nausea | 33/16.5 | 167/83.5 |  | 0.076 |
| Irregularities in menstruation | 0/0 | 200/100 |  | 0.420 |

Chi-square test for correlation analysis with $\chi^2 = 0.39832$ (age), 0.1564 (gender) at a significance level $P < 0.05$ .

**Table 4b:** Correlation between Previous Infection with Covid-19 and Adverse Events.

| Parameter | Infection with Covid-19 |  |  | P-value |
| --- | --- | --- | --- | --- |
|  | No<br>(count/percentage) | Yes<br>(count/percentage) | Total<br>(count/percentage) |  |
| Age: |  |  |  |  |
| Less than 20 | 29/61.7 | 19/38.3 | 48/100 | 0.9852 |
| 20 to 30 | 111/60.7 | 72/39.3 | 183/100 |  |
| 31 to 40 | 98/60.1 | 65/39.9 | 163/100 |  |
| 41to 50 | 39/59.1 | 27/40.9 | 66/100 |  |
| More than 50 | 31/64.6 | 17/35.4 | 48/100 |  |
| Total: | 308/60.6 | 200/39.4 | 508/100 |  |
| Gender: |  |  |  |  |
| Male | 210/59.4 | 67/38.8 | 277/100 | 0.6938 |
| Female | 98/61.2 | 133/40/6 | 231/100 |  |
| Total: | 308/60.6 | 200/39.4 | 508/100 |  |

Chi-square test for correlation analysis at a significance level $P < 0.05$ .

**Table 4c:** Correlation between Age/Gender and Adverse events due to vaccine:

|  | Injection-site pain |  | Headache |  | Fever |  | Fatigue |  | Muscle aches |  | Nausea |  | Irregularities in menstruation |  |
| --- | --- | --- | --- | --- | --- | --- | --- | --- | --- | --- | --- | --- | --- | --- |
|  | Yes | P-value | Yes | P-value | Yes | P-value | Yes | P-value | Yes | P-value | Yes | P-value | Yes | P-value |
| Age: |  |  |  |  |  |  |  |  |  |  |  |  |  |  |
| Less than 20 | 13 (2.6) |  | 11(2.2) |  | 12 (2.4) |  | 16 |  | 9 |  | 6 |  | 0 |  |
| 20 to 30 | 66 (13.0) |  | 60 (11.8) |  | 55 (10.8) |  | 63 |  | 49 |  | 25 |  | 1 |  |
| 31 to 40 | 26 (5.1) | <0.0001<br>**** | 25 (4.9) | <0.0001<br>**** | 23 (4.5) | <0.0001<br>**** | 29 | <0.0001<br>**** | 23 | 0.002** | 25 | 0.093 | 0 | 0.776 |
| 41 to 50 | 1 (0.2) |  | 6 (1.2) |  | 4 (0.8) |  | 9 |  | 4 |  | 2 |  | 0 |  |
| More than 50 | 8 (1.6) |  | 10 (2.0) |  | 10 (2.0) |  | 13 |  | 9 |  | 9 |  | 0 |  |
| Gender: |  |  |  |  |  |  |  |  |  |  |  |  |  |  |
| Male | 45 (8.9) |  | 38 (7.5) |  | 35 (6.9) |  | 50 |  | 35 |  | 22 |  | 0 |  |
| Female | 69 (13.6) | 0.070 | 74 (14.6) | 0.711 | 69 (13.6) | 0.774 | 80 | 0.091 | 59 | 0.276 | 45 | 0.947 | 1 | 0.149 |

Chi-square test for correlation analysis at a significance level \*P < 0.05, \*\* P < 0.01, \*\*\* P < 0.001, \*\*\*\* P < 0.0001.
The values are represented as count (percentage).

## Discussion

Covid-19 has had a tremendous social and economic influence on the entire planet. Rapid human-to-human transmission has created a significant public health risk [26]. Since the start of Covid-19 pandemic in 2020, different management approaches were developed and implemented to contain the disease. Vaccine development started in an attempt to provide sufficient immunization and maintain herd immunity. Yet, people have expressed their concern about the vaccines efficacy and safety which led to refusal or hesitancy to receive the vaccine [18]. There is a worldwide variation in people’s confidence in the vaccine which depend on several factors such as age, gender, and income in addition to the availability and accessibility of the vaccines [16, 17]. Moreover, social media has played an important role in influencing people’s perception of the vaccines by pointing out the importance of vaccination [24]. However, some information was misleading which resulted in a negative impact on people’s confidence in the vaccine. Certain adverse effects are unlikely to show in pre-licensure clinical research due to their low frequency, small number of participants, and other study constraints. Thus, post-marketing surveillance is crucial to ascertain the safety and effectiveness of the vaccine [27].

In our study, we evaluated the short-term adverse effects following vaccine administration. In addition, we investigated people’ perception and acceptance of the vaccine and determined the factors which can influence their decision making. Our study observed that the age of most of our participants was between 20 to 40 years old and majority were females. Most of the study population was from medical profession from Delta region, Egypt. The majority of our participant reported that risk factors associated with Covid-19 infection were chronic diseases such as cardiovascular diseases, followed by work environment, respiratory conditions, advanced age, autoimmune diseases, and cancer.

Consistently, people who perceive a higher risk of Covid-19 infection are more inclined to favor the vaccination [28]. The considerable proportion of younger responders with no medical disease contributes to the low percentage of perceived severity [29].

*Mohamed et al.* [29] study showed that participants reported difficulties to receiving the Covid-19 vaccination because of its side effects, vaccine availability, and alarming vaccine material on social media. The majority believed that Covid-19 immunization may protect them and others from illness. This is similar with other nations’ findings [18, 30, 31]. Furthermore, respondents considered that the vaccine was useful owing to Ministry of Health (MOH) recommendations and the fact that they could live a normal life after immunization [29].

In contrast, additional research is needed to assess vaccinations’ efficacy to prevent disease transmission. After receiving the Covid-19 vaccine, persons should continue to wear masks, wash their hands often, and exercise physical separation until herd immunity is obtained [28, 32]. Perceived susceptibility, benefit, and signals to action are related to better acceptance of the Covid-19 vaccination, which is consistent with prior research [33–35].

Our study found that 78.5% of the participants received Covid-19 vaccine and percentage of participants who are willing to take the vaccine about 43 % of participants who did not receive the vaccine. Previous research has found that being married, highly educated, experienced, or a doctor working in the healthcare setting are all connected with improved perception, attitude, and readiness to get Covid-19 vaccinations [18, 36–40], which is consistent with our findings. As these demographic factors may be associated with better levels of knowledge, responsibility, and care for family, which may lead to improved vaccination uptake.

The majority of our participants received adequate information about the vaccine and trusted the information provided. Workplace advice which might have influenced people’s perception have been studied as well.

Inconsistent with prior research [41, 42] found that notwithstanding the fact that the majority of participants obtained information from the MOH, readership of scientific papers, which are the source of evidence-based information, was relatively low.

According to the data of recent study, the MOH was the primary source of information regarding Covid-19 for almost three-quarters of the population [43]. Other studies have found that media and social media were the primary sources of information about Covid-19 for the general public [28, 44, 45].

Inadequate vaccination knowledge can be attributed to a lack of education, a low socioeconomic standing, or information obtained from peers [46, 47]. According to this survey, more than half of the respondents were unaware about the Covid-19 vaccination.

Higher knowledge scores were substantially related with higher education level, higher wealth, and living with high-risk persons. Malaysians were found to have high levels of knowledge, attitude, and perception on Covid-19 prevention [47]. This is perhaps the key explanation for respondents’ increased acceptance of Covid-19 vaccination despite their lack of understanding about the vaccine. Our acceptance rate is approximately similar to that of Saudi Arabia (64.7%) [48] and the United Kingdom (64%) [49], higher than that of Turkey (49.7%) [50], but lower than that of China (91.3%) [36] and Indonesia (93.3%) [28]. However, as compared to the recent study *by Shawki et al.* [23] our study provided higher acceptance rates (76% *vs.* 14%). Acceptance of the Covid-19 vaccination was substantially linked with being younger, having a better education level, being female, and not having chronic conditions. In Saudi Arabia, older age groups, married people, those with a postgraduate degree or higher, non-Saudis, and those working in the government sector were more likely to receive the Covid-19 vaccination [48].

Although the acceptance percentage is comparable to that published studies, one significant difference is that most of our participants are middle aged, whilst in Malaysia [29], younger age groups were more accepting, older age groups were more accepting in Saudi [48].

The main reasons were the lack of sufficient studies that provide evidence of safety and effectiveness (64%) and fear of adverse events (61%), while those having contraindication to the vaccines were only 21% of all respondents.

The risk of Covid-19 illness was the most common reason for acceptance among people who elected to take the vaccination after it became available[51]. While the most prevalent cause for vaccine refusal and reluctance was a lack of awareness about its safety and a lack of appropriate clinical studies. This was consistent with virtually all research conducted to investigate Covid-19 vaccine reluctance [52, 53].

Data received from participants in this trial demonstrated that adverse effects of the vaccination were documented after two doses, with the majority happening after the second dosage. Injection site soreness, headaches, flu symptoms, fever, and weariness are the most prevalent symptoms. A rapid pulse, generalized pains, trouble breathing, joint discomfort, cold, and sleepiness are less typical adverse effects. Bell’s palsy and lymph node soreness and edema were rare adverse effects.

These findings were consistent with the FDA Fact Sheet for Recipients and Caregivers, which stated that influenza-like symptoms were more prevalent in those under 60, but discomfort at the injection site was more common in people 60 and older [54]. The most frequent side responses, which include discomfort at the site of administration, weariness, headaches, muscle and joint soreness, chills, and raised body temperature, might linger for days, according to the information sheet, and were more likely after the second injection than the first [54].

According to *El-Shitany et al.,* [55] study, the injection site discomfort was reported more frequently among people aged 60 and over (80.8%) than among younger people (68.6%). A relatively low number of people reported injection-site edema and redness. Following the second dosage, the percentage of those experiencing local symptoms climbed dramatically. Contradict to *Polack et al.* [56], who found that injection site discomfort was more common in those under the age of 55 compared to those over the age of 55.

Our findings are consistent with those of *Polack et al.* [56], who found that vaccination-associated systemic adverse effects were more prevalent in younger persons and more common after the second vaccine dosage. According to the two studies [55, 56], headaches were the most prevalent complaint following a vaccination injection. In addition, fever was more likely among young person’s following the second dosage.

While vaccinations are being prepared, ongoing education should be provided to promote knowledge and clear up any misunderstandings or misinformation concerning the vaccine. Health education should ideally be comprehensive, multilingual, and accessible to laypeople. The critical messages should reach all residents from all walks of life, including those in rural areas and those who are technologically illiterate. Printed materials and face-to-face public presentations, in addition to web-based and application-based instructional tools, may help particular sectors of the population. Experts can hold public discussions involving religious organizations in places of worship.

The limitations of this study were that a cross-sectional design cannot determine the causal influence on outcomes. Data was collected via questionnaires, which are prone to over- and under-reporting. The use of convenience sampling via social media sites was one of the study’s limitations. Because most respondents were internet-savvy young individuals, the distribution of respondents may not reflect the real population. We propose a broader research with respondents from various origins, ethnicities, socioeconomic statuses, and regions. To improve the rate of response, many public platform sharing is required. Telephone interviews and face-to-face interviews, among other approaches, should be used to collect data.

The modest number of research participants who had previously been infected with the coronavirus suggests that additional investigations with an appropriate and compatible sample size for both previously infected and non-infected people are needed.

## Conclusions

This study showed that the participants generally had a good perception and acceptance of Covid-19 vaccination, which may be explained by access to factual information from trusted sources, mainly the ministry of health. Moreover, age, gender, and previous infection with Covid-19 significantly influenced tolerability of Covid-19 vaccine among participants. These findings imply that demographic factors impact how people view and perceive Covid-19 vaccinations, highlighting the need for further studies to inform strategies to fight against Covid-19. Awareness campaigns targeting all population must be maintained in order to deliver all updates.

## Declarations

### Ethical Approval

This study was conducted in accordance with the declaration of Helsinki and registered on ClinicalTrial.gov with code no.: NCT05857176. Institutional review board (IRB) approval is deemed unnecessary for online survey according to national regulations (Ref No.422CPD15).

### Consent to participate

Prior to commencing the questionnaire, informed consent was sought electronically using the form as this survey was entirely voluntary and anonymous.

#### Consent for publication

Not applicable.

## Data Availability

All relevant data is available in the present manuscript. The data presented in this study is available upon reasonable request from the corresponding author.

## Acknowledgment

The authors would like to extend their heartfelt gratitude to all of the participants who contributed to this study. Their openness to share their experiences and perspectives was invaluable in completing this research.

## Authors Contribution

R.H.W: Conceptualization, Investigation, designing of the study, Writing - final Draft, Review & Editing and Supervision. G.D: Conceptualization, Investigation, designing of the study, performing statistics, Writing - Original Draft, Review & Editing. M.A: extraction of data from the excel sheets, preparation of figures & tables and writing parts of the result section. M.Y: extraction of data from the excel sheets and writing part of the introduction. N.B writing part of the introduction and the results section. All authors have given approval to the final version of the manuscript.

