## Supplementary Table 1 for "Assessment of Safety and Acceptance toward Covid-19 Vaccines: A cross-sectional Study in Egypt"

**Supplementary Table 1: The English Version of used questionnaire in the study.**

| |
|---|
| Age |
| Gender |
| Occupation |
| Have you ever been diagnosed with COVID-19 infection? |
| Do you think COVID-19 infections are serious? |
| Is there had direct contact with COVID-19 patients? |
| Place of direct contact with patients. |
| Do you think you have risk factors that increase your chances of COVID-19 infection? |
| Risk factors that increase your chance to be infected by COVID-19<br>Advanced age<br>Autoimmune disease<br>Chronic disease<br>Respiratory conditions<br>Cancer<br>Work environment |
| Did you take COVID-19 vaccination? |
| If the answer is no, Do you intend to get a COVID-19 vaccination soon? |
| What types of COVID-19 Vaccine you received? |
| Do you believe in COVID-19 vaccines safety and effectiveness? |
| Did you receive enough information about COVID-19 ? |
| Do you think the information provided on COVID-19 vaccination enough? |
| Did you get advice from your workplace to receive COVID-19 vaccination? |
| What are the sources of your information about COVID-19 infection? |
| Did you will advise other to receive COVID-19 vaccination? |
| Why did you accept to take COVID-19 vaccination? |
| What the reasons why you hesitate or refuse to vaccinate COVID-19? |
| What factors can increase people's acceptance of COVID-19 vaccination? |
| What factors can increase people's acceptance of COVID-19 vaccination? |
| Adverse Effects Reported after Receiving Different Types of Vaccines<br>• Injection Site Pain |

| |
|---|
| • Headache<br>• Fever<br>• Fatigue<br>• Muscle aches<br>• Nausea<br>• Irregularities in menstruation |
| Did you infected with covid-19 virus after taking the vaccine? |
